# Prescription Trends of Initial Antihypertensive Medications among Treatment-Naïve Individuals in South Korea: A Retrospective Analysis

**DOI:** 10.1101/2024.04.08.24305523

**Authors:** Hajung Joo, Seung Eun Chae, Yeong Rok Eom, Nam Kyung Je

## Abstract

**Introduction:** Managing hypertension effectively is crucial for preventing cardiovascular complications. This study investigates the prescribing trends of initial antihypertensive treatments in South Korea, focusing on the shift towards combination therapies, especially fixed-dose combinations, as recommended by current guidelines.

**Methods:** We retrospectively analyzed data from 59,950 treatment-naïve hypertension patients without significant comorbidities from the National Patient Sample of the Health Insurance Review and Assessment Service, spanning 2009 to 2020. The study examined the prevalence of monotherapy versus combination therapy and the preferred classes of antihypertensive drugs.

**Results:** Among the cohort, 55.7% received monotherapy, and 44.3% were initiated on combination therapy. There was a notable increase in the prescription of fixed-dose combinations from 73.8% in 2009 to 93.8% in 2020. ARBs and CCBs were the predominant choices, with a preference for ARBs over ACE inhibitors, possibly due to the lower risk of side effects such as cough. The trend towards fixed-dose combinations aligns with guidelines advocating for improved patient adherence and efficacy.

**Conclusions:** The evolving prescription trends in South Korea towards combination therapy, particularly fixed-dose combinations, reflect a commitment to aligning with international hypertension management guidelines. This adaptation underscores the importance of evidence-based practice in enhancing hypertension care.

## Introduction

Cardiovascular disease (CVD) continues to be a major global health challenge. Hypertension, a key preventable risk factor, plays a pivotal role as a preventable risk factor for both CVD and mortality. ^1^ Despite the potential for control, poorly managed hypertension remains a growing health concern. ^2^ Effective management through appropriate drug therapy is crucial for reducing related complications and mortality.

Over the past 50 years, since the first hypertension guideline by the Joint National Committee (JNC) in 1977, ^3^ there has been significant progress in developing guidelines. These efforts aim to refine prescription practices through evidence-based review and provide valuable insights into appropriate drug selection. ^4^

With a successful start off with thiazide diuretics in the 1950s^5,6^, today, a robust line-up of ACE inhibitors (ACEI), angiotensin receptor blockers (ARB), calcium channel blockers (CCB) and thiazide/thiazide-like diuretics stands strong as first-line antihypertensives, as agreed by guideline consensus. ^7–9^

The management of newly diagnosed hypertensive patients typically involves monotherapy or combination therapy. ^7–9^ However, monotherapy often fails to achieve target blood pressure, underscoring the importance of combination therapy for improved cardiovascular outcomes. ^10–12^

Recent guidelines strongly recommend combination therapy as the first-line treatment for newly diagnosed patients. ^7–9^ Specifically, the 2017 American College of Cardiology/American Heart Association (ACC/AHA) guideline and the 2018 Korean Society of Hypertension (KSH) guideline recommend combination therapy as the initial treatment for newly diagnosed patients whose average blood pressure is more than 20/10 mmHg higher than the target blood pressure. ^7,8^ Moreover, the European Society of Cardiology and the European Society of Hypertension (ESC/ESH) guideline takes a more proactive approach, recommending the initiation of treatment with combination therapy for most patients, with a few exceptions. ^9^ This combination therapy is available in two forms: fixed-dose combination (two or more drugs combined in a single-pill at predetermined doses) or free-dose combination (two or more drugs taken separately at flexible doses). Further looking at combination therapy, the 2018 ESH guideline suggests initiating treatment with a two-drug combination, preferably a fixed-dose combination. For patients with uncomplicated hypertension, the recommendation is to use either an ACEI or ARB with CCB or thiazide diuretics. In a broader sense, the ACC/AHA guideline advises the use of two first-line agents from different drug classes, and the KSH guideline provides a general recommendation, allowing the use of two drugs in combination. ^7–9^ Despite varying approaches, these guidelines converge in favoring fixed-dose combination therapy over free-dose combination, as it is believed to maximize blood pressure reduction and enhance medication adherence. ^7–9,13^

Given these evolving guidelines and the diversity in first-line drug class and agent selection, it is crucial to examine the current prescription practices in antihypertensive drug therapies, especially in the context of treatment-naïve patients. ^4^ While previous studies have explored these, there is a lack of updated information post the release of the 2017 and 2018 guidelines. ^14–17^ Hence, our study aimed to analyze prescription patterns and trends of antihypertensive drugs in treatment-naïve patients with uncomplicated hypertension from 2009 to 2020 in South Korea.

## Methods

### Data Source

The data used for analysis was drawn from the HIRA-NPS (National Patient Sample), which is a random sample of patients extracted from the claims data by the Health Insurance Review and Assessment Service (HIRA), with the percentage being 3% before 2019 and 2% thereafter. ^16^ To match consistency with the 3% data, the percentage was adjusted in the data table for years 2019 and 2020. This dataset, updated with a different set of patients annually, includes patient demographics such as age and sex, diagnoses, types of healthcare institutions, region, and provider demographics. We conducted an analysis for the period 2009 – 2020.

The diagnostic codes were sourced from the KCD, 5-7^th^ edition (KCD-5,6,7). Drug codes were obtained from the HIRA Reimbursement Drug List (2009-2020).

### Study Population

To identify a cohort of patients with treatment-naïve, uncomplicated hypertension, this study analyzed HIRA-NPS data. It included patients aged 20 years or older who were newly diagnosed with essential hypertension (KCD 5-7 code I10) between July 1 – Dec. 31 of each year and who had no compelling comorbidities.

The index date was set as the day patients received their first prescription of the antihypertensive agent during this period. To ensure that it is their first diagnosis of hypertension, patients were excluded if they had any history of hypertension diagnosis and/or any antihypertensive drugs (even if for another indication) in period 6 months prior to index date. Patients with history of cardiovascular comorbidities such as complicated hypertension (I11-I15), atrial fibrillation (I48), heart failure (I50), ischemic heart disease including myocardial infarction (I21-I25), peripheral vascular disease (I70, I73), transient ischemic attack (G45), angina (I20), cerebrovascular disease including stroke (I63-I64), renal failure (N03-N05, N17-N19, Z49, Z94.0, Z99.2), and diabetes (E11-E14), within the 6 months before the index date were also excluded. This exclusion aimed to ensure the selection of treatment-naïve patients and reduce the likelihood of including individuals with pre-existing hypertension, as the presence of comorbidities can increase the chances. Patients whose record did not include the hypertension-related condition in the 6-month period before the index date were considered treatment-naïve, uncomplicated hypertension patients. The patients in the final cohort of uncomplicated hypertension patients were excluded if there was no record of receiving antihypertensive drug treatment (Figure 1).

**Figure 1.**
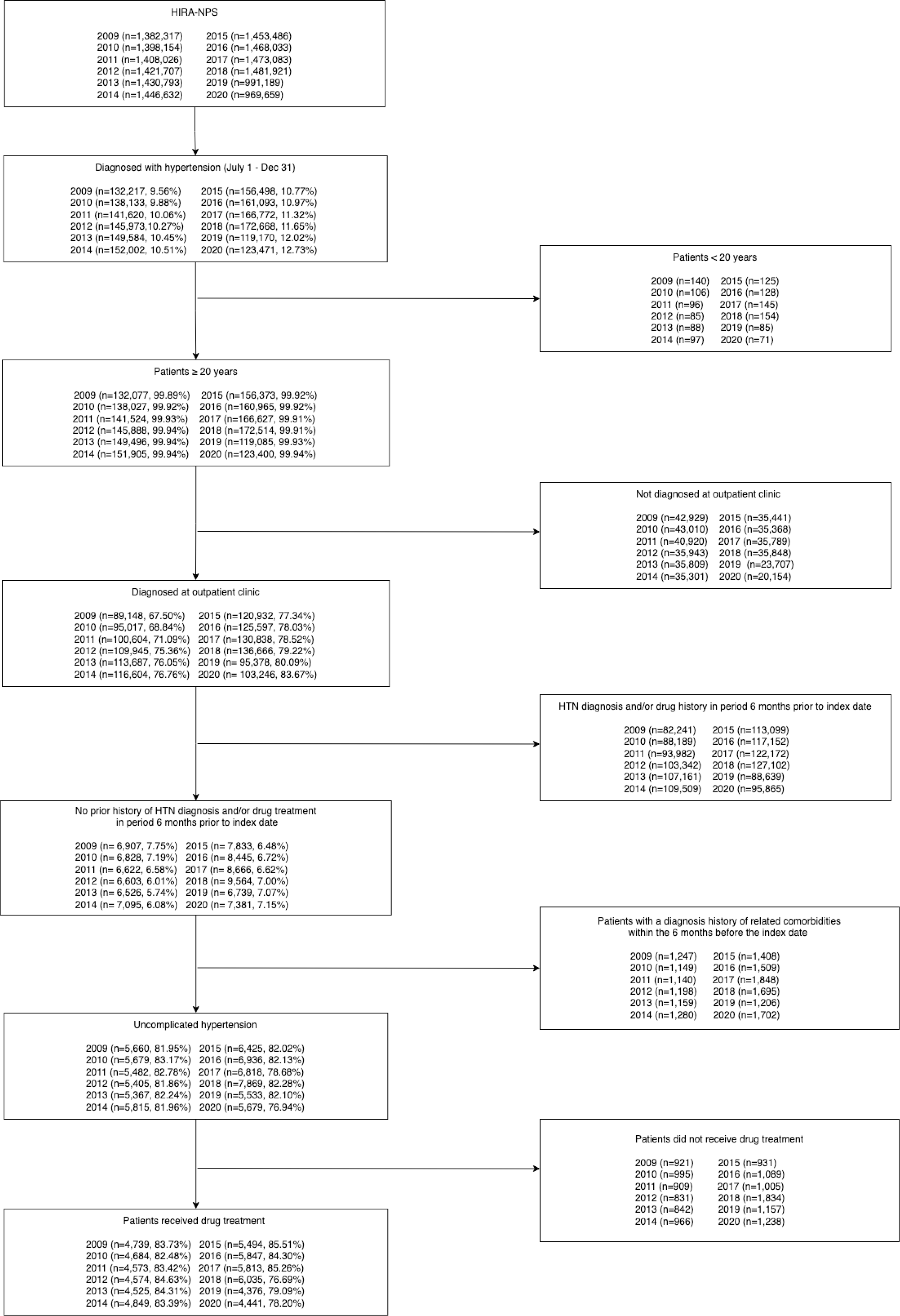
Study population flow chart

The study population was categorized into five age groups (20-39, 40-49, 50-59, 60-69, and 70+), three regions (capital, metropolitan, and other), and three clinic specialties (general practice, internal medicine, and other).

Comorbidities were defined by KCD codes, 5-8^th^ edition: dyslipidemia (E78), cancer (C00 – C99), gout/hyperuricemia (M10, E79.0), osteoporosis (M80-M82), and benign prostate hyperplasia (BPH) (N40).

### Antihypertensive Drugs

Antihypertensive agents were categorized into six groups: ARB, ACEI, CCB, beta-blockers (BB), thiazide or thiazide-like diuretics, and others (including alpha-blockers, loop diuretics, and potassium-sparing diuretics).

First-line drug treatment regimens were primarily divided into two types: monotherapy and combination therapy. Monotherapy involved the use of a single type of antihypertensive drug, while combination therapy incorporated two or more distinct drugs from different classes. Combination therapy was further divided into fixed-dose (single-pill) or free-dose combinations (separate pills). The list of drugs and combinations is provided in the supplementary table (Supplementary Table 1). Patients who had records of both monotherapy and combination therapy prescriptions were categorized as part of the combination therapy group.

Regarding antidyslipidemic and antihypertensive fixed-dose combination medications (i.e., atorvastatin/amlodipine, atorvastatin/irbesartan), only the antihypertensive drugs were considered as part of the monotherapy regimen.

### Statistical Analysis

This cross-sectional study aimed to assess the factors influencing the selection of first-line drug therapy in treatment-naïve patients with uncomplicated hypertension who were newly prescribed antihypertensive medications from 2009 to 2020. Descriptive analyses were conducted to describe the baseline characteristics of patients using counts and percentages. The chi-squared test was utilized to determine the statistical significance of patient characteristics associated with the prescribing trends, including 1) monotherapy vs combination therapy, 2) fixed-dose combination vs free-dose combination in combination therapy, and 3) antihypertensive regimen. The data were also presented in counts and percentages. Multiple logistic regression analysis was employed to identify influential factors in combination therapy prescriptions and fixed-dose combinations by estimating odds ratio (OR) with a 95% confidence interval. We further tracked the annual trend and examined the changing popularity of both drugs and drug classes. The Cochran-Armitage test was applied to assess the linear time trends of drug therapies throughout the study period.

All analyses were performed using the R software (version 3.5.1; R Foundation for Statistical Computing, Vienna, Austria). A P-value of <0.05 was considered statistically significant.

## Results

### Baseline Characteristics

We analyzed 59,950 patients with uncomplicated hypertension who began drug treatment. Of these patients, 58.3% were male. The most represented age group was 50-59 years, accounting for 33.0% of the total. From 2009 to 2020, the proportions of treatment-naïve patients increased from 7.9% to 11.1% (adjusted for comparison). Within the patient cohort, there were individuals with comorbidities, including dyslipidemia (21.3%), osteoporosis (4.1%), cancer (3.1%), benign prostatic hyperplasia (BPH) (2.7%), and gout/hyperuricemia (2.5%). Furthermore, 53.0% of the patient group resided in non-capital and non-metropolitan areas, and 51.8% of the prescriptions were from internal medicine specialists (Table 1).

**Table 1.**
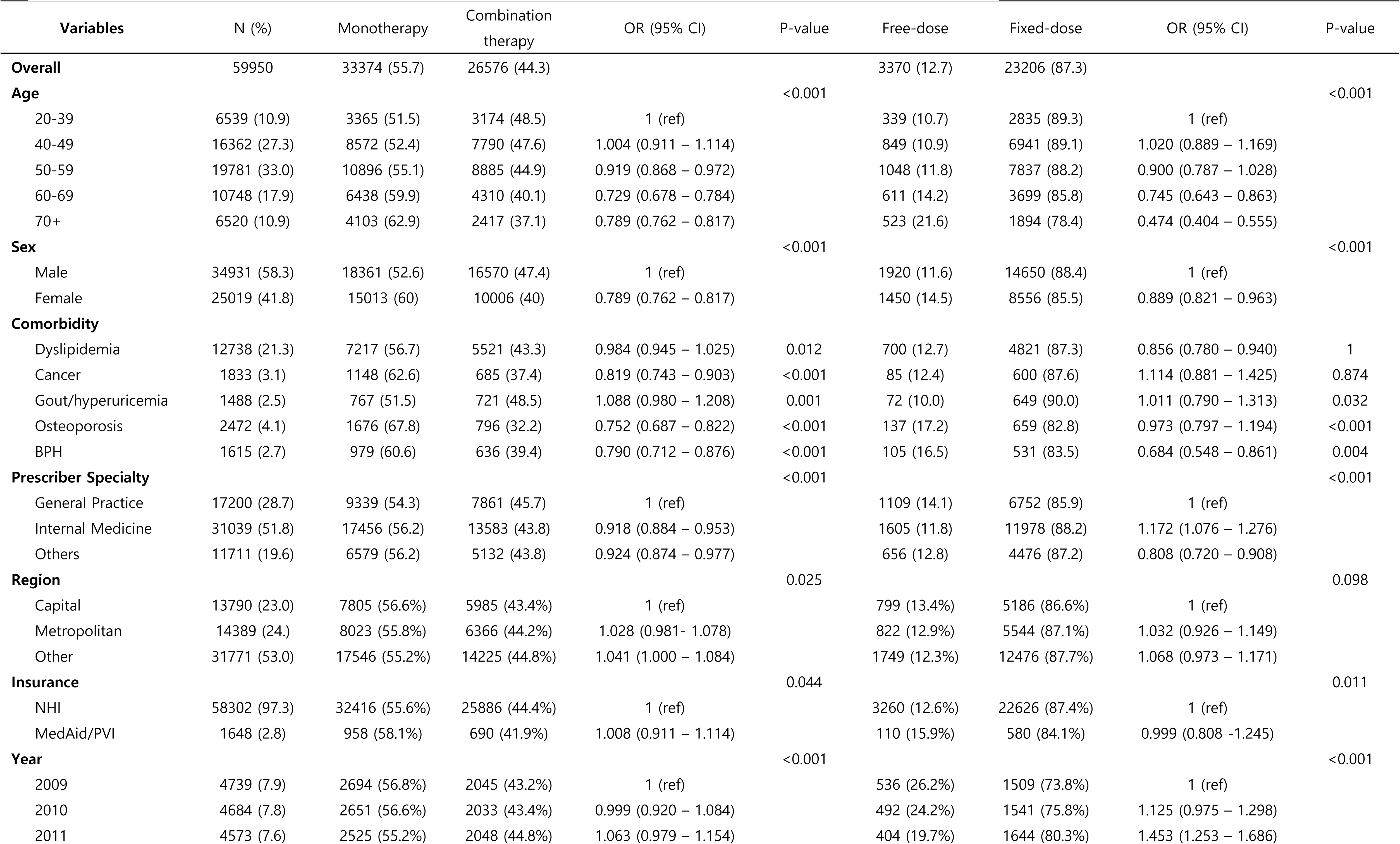
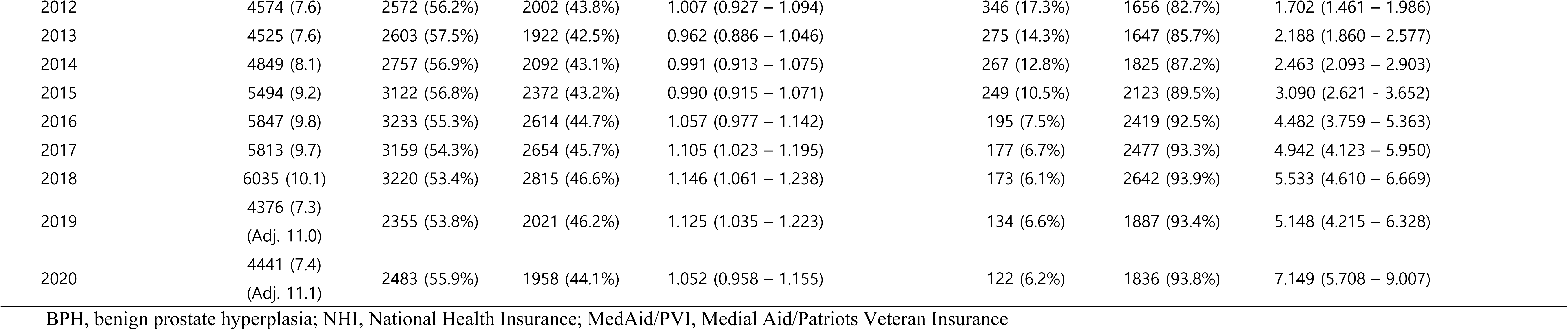
Baseline characteristics and factors associated with prescription pattern by drug regimen.

### Trends Associated Initial Antihypertensive Drug Treatment Regimen

During the study period, a higher proportion of patients received monotherapy (55.7%) compared to combination therapy (44.3%). Among those prescribed combination therapy, the majority (87.3%) were given fixed-dose combinations, in contrast to free-dose combinations (12.7%). The preference for fixed-dose combinations significantly increased from 73.8% to 93.8%, while the use of free-dose combinations correspondingly declined from 26.2% to 6.2% (Cochran–Armitage Trend (CAT) test; *p* < 0.001) as illustrated in Figure 2A.

**Figure 2.**
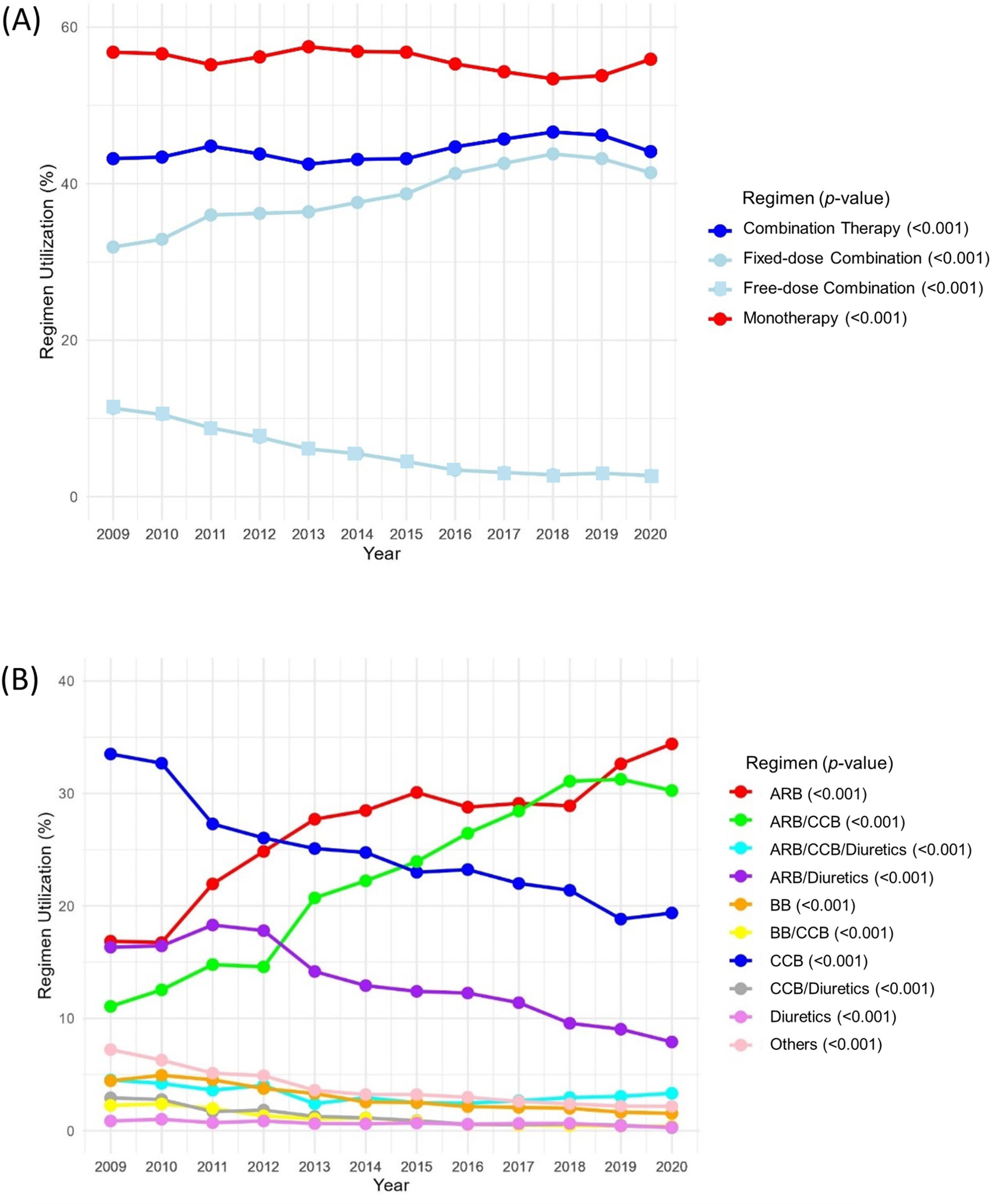
Prescription trends of initial treatment (A) Monotherapy vs. combination therapy (B) Drug regimen ARB, angiotensin receptor blockers; BB, beta-blockers; CCB, calcium channel blockers Statistical significance was determined using p-values from the Cochran-Armitage trend test

Regarding specific regimens, the most commonly prescribed were ARB, CCB, ARB/CCB, and ARB/diuretic. Notably, the use of ARB and ARB/CCB regimens increased significantly, from 16.9% to 34.4% (CAT test; *p* < 0.001) and 11.1% to 30.3% (CAT test; *p* < 0.001), respectively. In contrast, the usage of CCB and ARB/diuretic declined from 33.5% to 19.4% and 11.1% to 8.9%, respectively. Initially, CCB was the most prescribed antihypertensive regimen (33.5% in 2009), but its usage decreased to 19.4% by 2020 (CAT test; *p* < 0.001). Meanwhile, ARB usage more than doubled from 16.9% in 2009 to 34.4% in 2020 (CAT test; *p* < 0.001). Although ARB/diuretic was initially the preferred combination therapy, it was later surpassed by ARB/CCB (Figure 2B).

In the ARB group, losartan was initially the most commonly prescribed drug. However, its usage declined over time, eventually being surpassed by telmisartan in 2020 (23.6% vs 24.0%). In the CCB group, amlodipine consistently remained the most prescribed drug, with its usage increasing from 66.4% in 2009 to 77.6% in 2020. The drug combinations within the ARB/CCB group varied over time; losartan/amlodipine was the predominant regimen in 2009 at 53.6%, but decreased significantly to 11.7% in 2020, with telmisartan/amlodipine becoming the most utilized combination in 2020. The ARB/diuretic group exhibited a similar pattern to the ARB group. Losartan/HCTZ was the most prescribed in 2009, but its usage declined overtime, being replaced by telmisartan/HCTZ, although not surpassed in 2020. Supplementary figure 1 illustrates these medication trends over the study period.

### Factors Associated with Initial Drug Regimen: Monotherapy vs Combination Therapy

The likelihood of receiving combination therapy over monotherapy decreased as age increased: 50 – 59 (0.919, 95% CI 0.868 – 0.972), 60 – 69 (0.784, 95% CI 0.736 – 0.835), and 70+ (0.729, 95% CI 0.678 – 0.784). Females were less likely than males to receive combination therapy (0.789, 95% CI 0.762 – 0.817). Patients with comorbidities such as cancer (0.819, 95% CI 0.743 – 0.903), osteoporosis (0.752, 95% CI 0.687 – 0.822), and BPH (0.790, 95% CI 0.712 – 0.876) also had a lower likelihood of receiving combination therapy. The odds of receiving combination therapy were higher when prescribed by a general practitioner compared to specialists (0.918, 95% CI 0.884 – 0.953 and 0.924, 95% CI 0.874 – 0.977) (Table 1).

### Factors Associated with Initial Combination Drug Regimen: Free-dose vs Fixed-dose

Among patients who receiving combination therapy, the likelihood of receiving fixed-dose combinations were lower in the 60 – 69 and 70+ age groups (0.745, 95% CI 0.643 – 0.863 and 0.474, 95% CI 0.404 – 0.555, respectively) compared to the 20 – 39 age group. Females were also less likely to receive fixed-dose combinations than males (0.889, 95% CI 0.821 – 0.963). Patients with dyslipidemia (0.856, 95% CI 0.780 – 0.940) or BPH (0.684, 95% CI 0.548 – 0.861) were less likely to receive fixed-dose combinations than those without these comorbidities. Internal medicine specialists were more likely to prescribe fixed-dose combinations (1.172, 95% CI 1.076 – 1.276), while other specialists were more likely to prescribe in free-dose combinations (0.808, 95% CI 0.720 – 0.908), compared to general practitioners. Nevertheless, a notable preference of fixed-dose combinations was illustrated by their consistent increase over the years from 2009 to 2020 (7.149, 95% CI 5.708 – 9.007)

## Discussion

The growing concern over the high prevalence of hypertension and the consequences of leaving it untreated and uncontrolled have led recent guidelines to continually evolve in their recommendations for the initial treatment of hypertension. To shed light on this evolving landscape, our study analyzed prescribing trends in the use of antihypertensive drugs and treatment patterns in treatment-naïve, uncomplicated hypertensive patients from 2009 to 2020, utilizing HIRA-NPS claims data.

Recent clinical guidelines recommend the adoption of tight initial treatment approaches for hypertension to reduce the risk of cardiovascular complications in most hypertensive patients. In line with these guideline recommendations, our data reveals that in South Korea, while monotherapy remained more prevalent in practice compared to combination therapy, the increasing trend of fixed-dose combinations was evident through a continuous rise over the study period. This shift is driven not only by the cost-effectiveness of a single pill but also by improved medication adherence, as patients need to take only one pill instead of multiple. This positive development is noteworthy in the treatment of hypertension as the enhanced adherence through simplified regimen offered by fixed-dose combinations align with the goal of improving patient compliance, ultimately leading to better blood pressure control and reducing the risk of cardiovascular events. Additionally, the widespread availability of fixed-dose combination drugs in the market has contributed to the increasing trend in prescription rates. ^18^ The upward trajectory of fixed-dose combinations underscores a progressive approach in addressing the complexities of multiple conditions, marking a significant stride in optimizing therapeutic strategies for improved patient outcomes.

There has been a noticeable shift in preference when analyzing yearly trends in the most commonly prescribed drug regimens. In monotherapy, the usage of ARB increased (from 16.9% to 34.4%), while the usage of CCB decreased (from 33.5% to 19.4%), along with other monotherapy regimens. Additionally, the contrasting rarity in the prescription of ACEI underscores the pronounced preference for ARB in South Korea. While, ACEI remains a common prescription choice in the US^19^ and the UK^20^. Despite both ARB and ACEI being renin-angiotensin-aldosterone system (RAAS) inhibitors, the preference for ARB over ACEI in Korea can be attributed to the higher prevalence of a side effect associated with ACEI - dry cough. Previous research indicates that Asian populations may be more susceptible to experiencing the side effect of dry cough when taking ACEI. ^21,22^ Consequently, the availability of an alternative drug class, ARB, which is also RAAS inhibitors with a more favorable side effect profile, might have contributed to the low usage rate of ACEI.

In combination therapy, the utilization of the ARB/diuretic combination has decreased since 2011, paving the way for the rise of ARB/CCB combination as the preferred choice, starting in 2009 (Table 2). This shift may be influenced by compelling findings, such as those from the ACCOMPLISH clinical trial, which favored ACEI/CCB over ACEI/diuretic in 2008. ^23^ Since the first JNC guideline in 1977, ^24^ diuretics played a pivotal role as the first-line treatment until its 7th edition in 2003. ^25^ However, over time, a shift in preference becomes evident, with ARB and CCB gaining prominence among antihypertensive drug classes. This trend holds true for previous studies conducted in South Korea. ^14,26^ The introduction of fixed-dose combinations encompassing these two drug classes, ARB and CCB, further solidified this preference in clinical practice. However, in the US^27,28^ and Japan^29^, ARB/diuretic combinations were favored compared to ARB/CCB combinations.

Diving into the ARB/CCB combination drugs, our study identified valsartan/amlodipine as the leading medication in combination therapy, but this was later replaced by telmisartan/amlodipine in 2018. The decline in valsartan usage and the subsequent increase in telmisartan can be primarily attributed to a valsartan recalls and the associated safety concerns. On July 5, 2018, the European Medical Agency issued a recall due to the detection of N-Nitrosodimethylamine (NDMA), a carcinogenic substance, in the active pharmaceutical ingredient of valsartan supplied by Zhejiang Huahai Pharmaceuticals. ^30^ This recall raised widespread concerns among prescribers and patients, prompting them to seek safer alternatives. In our study, telmisartan, another ARB drug, emerged as a viable and perceived safer option in response to the valsartan contamination issue. This transition underscores the critical role that safety and regulatory actions can play in reshaping drug utilization trends.

Our study identified disparities in factors related to the use of antihypertensive drugs. One observation pertains to gender-based differences in the choice of initial antihypertensive drug regimens. Our research highlights that females are less likely than males to initiate hypertension treatment with combination therapy. This observation aligns with existing knowledge of gender differences in hypertension control rates, prevalence, and awareness. ^31–33^ Research conducted in South Korea further illustrates this trend, revealing that hypertension is more common in men, while its prevalence comparatively increases with age in women^34^. While biological factors may play a role in this disparity, the conservative approach to initial treatment, as evidenced by the lower rate of combination therapy among females – despite it being the recommended approach - echoes concerns about lower disease control rates among women. ^33,35–37^ The global objective remains focused on achieving timely blood pressure control in hypertension to reduce the risk of developing cardiovascular disease. ^38^ Therefore, it is worth to consider and address such gender-based differences in future discussions, noting that current guidelines do not include gender-specific recommendations.

We also observed that older age groups were less likely to receive combination therapy and, furthermore, less likely to receive fixed-dose combinations as their initial treatment. The management of hypertension in older adults has been a topic of debate and discussion, with concerns such as orthostatic hypotension, dose adjustment, multimorbidity, and frailtiy. ^39,40^ These prevalent concerns may contribute to the observed conservative treatment approach among older age groups.

Consequently, prescribers may exercise caution in initiating aggressive drug treatment through a titration approach with low doses and monotherapy^41^ Nonetheless, in terms of monotherapy, CCB, recommended for elderly patients by guidelines, were the most commonly prescribed drug class among the 60-69 and 70+ age groups (29.8% and 36.5%, respectively), indicating adherence to guideline recommendations for drug class selection despite the lower utilization of fixed-dose combinations.

As a related consideration of disease-related therapy, patients with osteoporosis and benign prostatic hyperplasia (BPH) were also found to be less likely to receive combination therapy. Osteoporosis and BPH are both common age-related diseases that become more prevalent with increasing age. The 2017 ACC/AHA guidelines recommend alpha-blockers as a second-line agent for hypertensive patients with BPH. ^7^ While the use of the ‘Other’ regimen, which includes alpha-blockers, was highest in patients with concomitant BPH (6.2%), those with BPH were still less likely to receive combination therapy compared to those without this comorbidity. This trend may be attributed to the age-related nature of the disease, which aligns with the lower usage of combination therapy in older patients. Additionally, for patients with BPH, we observed that fixed-dose combinations were also less likely to be prescribed. Currently, there are no available fixed-dose combination drugs with alpha-blockers, which may contribute to the low usage of fixed-dose combination therapy in patients with concomitant BPH who are prescribed alpha blockers.

Our study comes with limitations to consider for careful interpretation of our findings. Firstly, our reliance on claims data initially collected for reimbursement purposes raises the possibility of inaccurate diagnostic information. Additionally, the claims data lack clinical information, such as blood pressure measurements, contraindications influencing the choice of antihypertensive drugs, and details on medication adherence, all of which significantly impact the selection of antihypertensive drug treatment. Secondly, although our study design allowed for the review of six months of patient history preceding the index date, our cross-sectional analysis, based on year-specific data, may not fully capture the continuum of patient experiences with hypertension. This limitation implies that our identification of patients with uncomplicated hypertension might not fully account for their historical clinical context. These limitations highlight the need for caution in generalizing our results and underscore areas for potential improvement in future research.

Nonetheless, our study presents a major strength in its utilization of HIRA-NPS dataset, which offers a significant advantage by providing a national representative sample of the South Korean population. The extensive scope of the dataset enables our findings to accurately reflect national trends and practices related to initial antihypertensive drug use. This national representation allows for more confident generalizations and applications to healthcare decision-making based on the observed patterns, as it minimizes the risk of selection bias.

## Conclusions

In conclusion, a higher proportion of treatment-naïve patients with uncomplicated hypertension received monotherapy as their initial drug treatment (55.7%), compared to combination therapy (44.3%). Within the context of combination therapy, fixed-dose combinations were predominantly favored over free-dose combinations. The preference for fixed-dose combinations was further confirmed by their consistent growth in usage from 2009 to 2020. Amlodipine was the most prescribed medication in monotherapy and valsartan/amlodipine was the most prescribed medication in combination therapy.

## Author declarations

### Ethical Approval

The Institutional Review Board of Pusan National University approved this study (PNU IRB/ 2023_131_HR).

### Consent to participate

Not applicable

### Consent to publish

Not applicable

### Authors’ contributions

HJ and NKJ conceived and designed the study; HJ took the lead in analyzing the data and drafted the initial manuscript.; All authors contributed to drafting the article and endorsed the final version for submission to publication.

### Data availability

HIRA-NPS data were used in this study and were not permitted to be shared. Raw data were obtained with permission from the Health Insurance Review and Assessment Service of Korea (http://opendata.hira.or.kr).

